# Using Artificial Intelligence to Learn Optimal Regimen Plan for Alzheimer’s Disease

**DOI:** 10.1101/2023.01.26.23285064

**Authors:** Kritib Bhattarai, Trisha Das, Yejin Kim, Yongbin Chen, Qiying Dai, Xiaoyang Li, Xiaoqian Jiang, Nansu Zong

## Abstract

**Background:** Alzheimer’s Disease (AD) is a progressive neurological disorder with no specific curative medications. While only a few medications are approved by FDA (i.e., donepezil, galantamine, rivastigmine, and memantine) to relieve symptoms (e.g., cognitive decline), sophisticated clinical skills are crucial to optimize the appropriate regimens given the multiple coexisting comorbidities in this patient population.

**Objective:** Here, we propose a study to leverage reinforcement learning (RL) to learn the clinicians’ decisions for AD patients based on the longitude records from Electronic Health Records (EHR).

**Methods:** In this study, we withdraw 1,736 patients fulfilling our criteria, from the Alzheimer’s Disease Neuroimaging Initiative(ADNI) database. We focused on the two most frequent concomitant diseases, depression, and hypertension, thus resulting in five main cohorts, 1) whole data, 2) AD-only, 3) AD-hypertension, 4) AD-depression, and 5) AD-hypertension-depression. We modeled the treatment learning into an RL problem by defining the three factors (i.e., states, action, and reward) in RL in multiple strategies, where a regression model and a decision tree are developed to generate states, six main medications extracted (i.e., no drugs, cholinesterase inhibitors, memantine, hypertension drugs, a combination of cholinesterase inhibitors and memantine, and supplements or other drugs) are for action, and Mini-Mental State Exam (MMSE) scores are for reward.

**Results:** Given the proper dataset, the RL model can generate an optimal policy (regimen plan) that outperforms the clinician’s treatment regimen. With the smallest data samples, the optimal-policy (i.e., policy iteration and Q-learning) gained a lesser reward than the clinician’s policy (mean -2.68 and -2.76 *vs*. -2.66, respectively), but it gained more reward once the data size increased (mean -3.56 and -2.48 *vs*. -3.57, respectively).

**Conclusions:** Our results highlight the potential of using RL to generate the optimal treatment based on the patients’ longitude records. Our work can lead the path toward the development of RL-based decision support systems which could facilitate the daily practice to manage Alzheimer’s disease with comorbidities.

## 1. Introduction

Alzheimer’s Disease (AD) is a progressive neurological disorder causing cognitive impairment and brain atrophy. Approximately 5.8 million people in the United States age 65 and older live with Alzheimer’s disease and approximately 60%-70% of 50 million people worldwide with dementia are estimated to be diagnosed with AD [1]. Currently, the etiology of AD is still unknown [2]. The scientific community has conflicting opinions about the cause of AD. Researchers have explored both neuroimaging and genetic paths to better understand the underlying causes of AD, yet there hasn’t been any conclusive outcome. Some researchers claim β-Amyloid plaque formation and aggregation causes AD [2] whereas others claim Apolipoprotein E (Apo E) gene along with various environmental factors do [3]; however, there is no consensus. Limited knowledge about AD’s pathogenesis has resulted in a wide range of speculations regarding the risk factors for AD, like vascular diseases, type-2 diabetes, traumatic brain injury, epilepsy, depression, smoking, diet, physical exercise, and alcohol consumption [4]. Due to the unknowns of AD’s etiology and risk factors, drug development has not made any significant progress and available drugs like cholinesterase inhibitors and memantine only treat the disease superficially. These drugs only help to temporarily ameliorate memory and thinking problems, but they do not treat the root cause of AD nor slow the rate of decline of a patient’s condition [5]. They are just aimed at modifying the disease symptoms only [6] [7].

Alzheimer’s disease management is further complicated by the high rate of comorbidities observed in patients [8]. Approximately 90% of AD patients are diagnosed with comorbid conditions [9]. Chronic diseases such as hypertension and depression are frequent among AD patients [10] [11]. Patients are mostly on several medications for other comorbidities. The relationship between AD and these comorbid conditions warrants further investigation on whether they act as risk factors or by-products of AD. Due to the limited knowledge of AD and its comorbid conditions, there is no consensus among physicians on how to manage such conditions [12]. This further complicates the management of AD. As a result, clinicians treat patients differently based on their own understanding and training experiences. Medication management ends up being a trial until a regimen temporarily relieves symptoms. As a result, it could take years of experience for a physician to medically manage AD with comorbidities [13]. A medication regimen learning tool can be very beneficial in this case to provide support to physicians. Such a learning tool would help physicians to learn the ways to properly treat AD patients with any possible comorbidities. For instance, a medication regimen learning tool could suggest a particular combination of drugs for each disease state for a patient instead of suggesting multiple combinations of drugs.

Furthermore, Artificial Intelligence (AI) has made it possible to create such medication regimen learning tools. It has been used to create such decision-support system models to predict drugs based on patient reviews [14]. Reinforcement Learning (RL) is an AI technology to learn a set of actions that can reward the most during the interaction of an agent in a specific environment (e.g., a computer game). Since RL is capable to learn a human-ish behavior and it achieved great success in diverse applications that require human interaction (e.g., Go [15]), there is a trend to adapt RL in healthcare (e.g., the regimen plan learned from Parkinson’s Disease [15] and Sepsis [16]). Such technology can learn from existing clinical data and provide suggestions to junior physicians who have less experience compared to senior physicians. This could revolutionize health care by transferring senior physicians’ years of experience to junior physicians through AI technologies. Here, we propose a study to learn the clinician’s treatment plan to facilitate the clinical practice of the junior clinician in managing AD patients. Specifically, we learn an RL-based model. It is a model based on stepwise decision-making and consists of states, actions, and rewards. A virtual agent checks the current state, explores different actions, and picks one that maximizes the future reward (Fig. 1). We have demonstrated that the proposed model outperforms the data-derived methods (e.g., transition probability-based model) for the patients with concomitant conditions (e.g., depression and hypertension). We came to this conclusion by comparing the Mini-Mental State Exam (MMSE) score from the data to the MMSE predicted by our RL model. The results highlight the proposed study to generate the clinician’s regimen plan for AD patients.

**Fig. 1.**
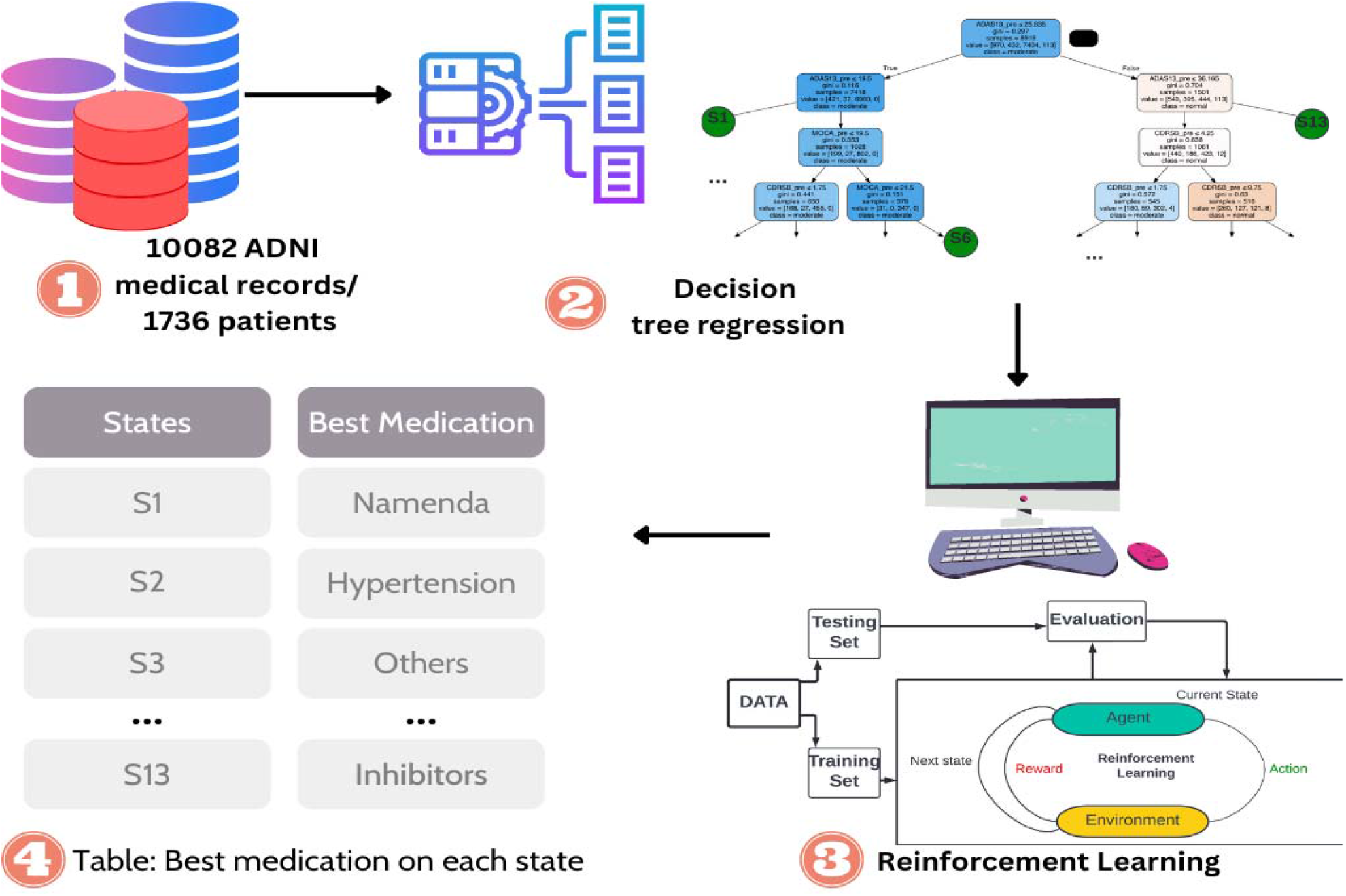
Pipeline of Reinforcement Learning-based regimen plan. (1) is the raw data that stores all the scores of lab tests like Alzheimer’s Disease Assessment Scale (ADAS), Montreal Cognitive Assessment (MoCA), Clinical Dementia Rating scale Sum of Boxes (CDRSB), Age, and so on. It also stores the medication applied and rewards based on Mini-Mental State Exam (MMSE) score. (2) 13 different states are defined using the decision tree. (3) A reinforcement learning model is prepared based on states from section (2) and actions and rewards from data in section (1). (4) Best medication/action is selected for each state after using reinforcement learning.

## 2. Methods And Materials

### 2.1. Data

The data is derived from Alzheimer’s Disease Neuroimaging Initiative (ADNI) database (adni.loni.usc.edu) which is the most frequently used open-access data in the pharmacogenomic studies for AD [17]. ADNI is a longitudinal multicenter study designed to support advances in AD prevention and treatment through the development of clinical, imaging, genetic, and biochemical biomarkers. One of the major goals of ADNI is to support advances in AD intervention, prevention, and treatment [18].

#### 2.1.1. Patient Cohort

We selected patients based on the following criteria: a minimum of two clinic visits, complete medical history, and clinical assessment data (Table 1). A total of 1,736 patients were selected (957 males and 779 females). Across all selected patients, the total number of visits was 10,082. The mean monthly visit per patient was 32.17 months and the mean number of visits per patient was 6.42 visits.

**Table 1.** Patient demographics for a different cohort of data (Whole Data, AD-only data, AD-hypertension data, AD-depression data, and AD-hypertension-depression data).

| Features | No. for Cohorts |  |  |  |  |
| --- | --- | --- | --- | --- | --- |
|  | Whole Data | AD-only | AD-Hypertension | AD-Depression | AD-Hypertension-Depression |
| No. of patients | 1,736 | 430 | 510 | 479 | 549 |
| No. of visits (s.d) | 6.42 (3.64) | 7.10 (3.65) | 7.35 (3.81) | 7.14 (3.66) | 7.35 (3.18) |
| Months of follow-up (s.d) | 32.17 (25.75) | 41.00 (26.89) | 43.22 (27.79) | 41.54 (26.87) | 43.46 (27.74) |
| Males | 957 | 257 | 305 | 283 | 324 |
| Females | 779 | 173 | 205 | 196 | 225 |
| Medications | Whole Data | AD-only | AD-Hypertension | AD-Depression | AD-Hypertension-Depression |
| Supplements/Others | 4,573 | 1,176 | 1502 | 1415 | 1701 |
| No drugs taken | 4500 | 1016 | 1246 | 1124 | 1337 |
| Cholinesterase inhibitors | 418 | 418 | 418 | 418 | 418 |
| Memantine | 176 | 176 | 176 | 176 | 176 |
| Anti-hypertensive drugs | 270 | 125 | 270 | 147 | 270 |
| Memantine-Cholinesterase inhibitors | 145 | 145 | 145 | 145 | 145 |

### 2.2. RL based Modeling

The traditional medical method of treating AD is assessing a patient’s current state and prescribing medication accordingly, then following up on the patient’s symptoms afterward. We utilized Reinforcement Learning (RL), a subfield of artificial intelligence (AI), to measure AD progression based on selected consecutive decisions. This consecutive decision-making nature of RL models is best described as a Markov decision process (MDP). An MDP consists of states, actions, and rewards where a state is Markovian if and only if the next state is dependent on the current state only. It is based on an agent at a certain state selecting different actions to maximize the rewards. The defined factors are described below.

*State s*: We define states as a finite set of a patient’s progression state in the latest clinic visit. Raw data on participants’ states were converted to discrete states. We picked up statistically significant features like Alzheimer’s Disease Assessment Scale (ADAS13) and age (Table 2) to predict the Mini-Mental State Exam (MMSE) score using regression. We then chose the significant variables and derived a decision tree (Fig. 2 and Table 2) to predict the MMSE scores. The decision tree divides each data into different ranges and then predicts the MMSE score. For example, an age of fewer than 70 years and an ADAS score of more than 20 could predict an MMSE score of 20. The predicted MMSE scores at the leaf nodes of a decision tree are our derived discrete states (Fig. 2). We grouped each visit according to the criteria specified by the decision tree and ignored states with less than 50 occurrences to avoid states without enough visits.

**Table 2.** Disease states classification based on a decision tree.

| Data Cohort | Disease State |  |  |  |  |  |  |  |
| --- | --- | --- | --- | --- | --- | --- | --- | --- |
|  | State | ADAS13 | RAVLT immediate | RAVLT Learning | Age | CDRSB | MOCA | FDG |
| Whole Data | S0 | ( ,19.5] | ( ,22.5] |  | ( ,77] |  |  |  |
|  | S1 | ( ,19.5] | ( ,22.5] |  | [78, ) |  |  |  |
|  | S2 | ( ,19.5] | (22.5, ) |  |  | ( ,1.8] |  |  |
|  | S3 | ( ,19.5] | (22.5, ) |  |  | (1.8, ] |  |  |
|  | S4 | (19.5, 25.8] |  |  |  | ( ,1.8] | ( ,19.5] |  |
|  | S5 | (19.5, 25.8] |  |  |  | (1.8, ] | ( ,19.5] |  |
|  | S6 | (19.5, 25.8] |  |  |  |  | (19.5, 21.5] |  |
|  | S7 | (19.5, 25.8] |  |  |  |  | (21.5, ] |  |
|  | S8 | (25.8, 36.2] |  |  |  | ( ,1.8] | ( ,19.5] |  |
|  | S9 | (25.8, 36.2] |  |  |  | (1.8, 4.2] | ( ,19.5] |  |
|  | S10 | (25.8, 36.2] |  |  |  | (4.2, ] | ( ,19.5] |  |
|  | S11 | (36.2, 42.7] |  |  |  |  |  |  |
|  | S12 | (42.7, ) |  |  |  |  |  |  |
| AD only | S13 | ( ,18.5] |  | ( ,1.5] |  |  |  |  |
|  | S14 | ( ,18.5] |  | (1.5, ) |  |  |  |  |
|  | S15 | (18.5, 25.2] |  |  |  |  | ( ,21.5] | ( ,6] |
|  | S16 | (18.5, 25.2] |  |  |  |  | ( ,21.5] | (6, ) |
|  | S17 | (18.5, 25.2] |  |  |  |  | (21.5, ) |  |
|  | S18 | (25.2, 35.8) |  |  |  | ( ,3.2] |  |  |
|  | S19 | (25.2, 35.8) |  |  |  | (3.2, ) |  | ( ,5] |
|  | S20 | (25.2, 35.8) |  |  |  | (3.2, ) |  | (5, ) |
|  | S21 | (35.8, ) |  |  |  |  |  |  |
| AD and Hypertension/<br>AD,<br>Hypertension<br>and Depression | S22 | ( ,17.5] |  |  |  |  | ( ,20.5] |  |
|  | S23 | (17.5, 22.2] |  |  |  |  | ( ,20.5] |  |
|  | S24 | ( ,22.2] |  |  |  |  | (20.5, ) |  |
|  | S25 | (22.2, 25.2] |  |  |  |  | ( ,13.5] |  |
|  | S26 | (22.2, 25.2] |  |  |  |  | (13.5, ) |  |
|  | S27 | (25.2, 31.2] |  |  |  | ( ,1.8] |  |  |
|  | S28 | (25.2, 31.2] |  |  |  | (1.8, 3.2] |  |  |
|  | S29 | (25.2, 31.2] |  |  |  | (3.2, ) |  |  |
|  | S30 | (31.2, 41.8] |  |  | ( ,75] |  |  |  |
|  | S31 | (31.2, 41.8] |  |  | (75, ) |  |  |  |
|  | S32 | (41.8, ) |  |  |  |  |  |  |
|  | S33 | (25.2, 31.2] |  |  |  | ( ,3.2] |  |  |
| AD and Depression | S34 | ( ,22.2] |  |  |  | ( ,3.75] | ( ,20.5] |  |
|  | S35 | ( ,22.2] |  |  |  | (3.75, ) | ( ,20.5] |  |
|  | S36 | ( ,22.2] |  |  |  |  | (20.5, ) |  |
|  | S37 | (22.2, 25.2] |  |  |  | ( ,3.75] | ( ,21.5] |  |
|  | S38 | (22.2, 25.2] |  |  |  | (3.75, ) | ( ,21.5] |  |
|  | S39 | (25.2, 35.8] |  |  |  | ( ,3.25] |  |  |
|  | S40 | (25.2, 35.8] |  |  |  | (3.25, ) |  |  |
|  | S41 | (35.8, ) |  |  |  |  | ( ,12.5] |  |
|  | S41 | (35.8, ) |  |  |  |  | (12.5, ) |  |

**Fig. 2.**
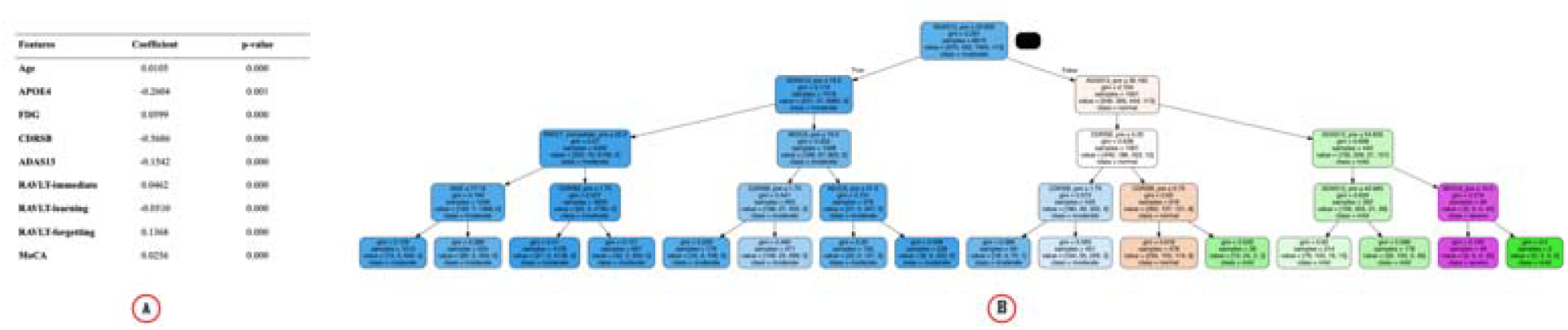
(A) Linear Regression table to find the statistically significant features. The significant features are then used for decision tree regression to classify data into different states. (B) Decision tree to predict MMSE scores.

*Action a*: We defined actions as a finite set of medications. Six combinations of drugs based on usage frequency were used: cholinesterase inhibitors (ChEIs), memantine, ChEIs +memantine, anti-hypertensive drugs, other supplements, and no drugs.

Hypertension drugs and other supplements are also include d to explore treatment across five data cohorts: Whole, AD-only, AD with hypertension, AD with depression, and AD with hypertension and depression. Please note that hypertension drugs and supplements are not traditional treatments for AD and are for patients with coexisting hypertension and other conditions [19–21].

*Reward r*: We defined reward as the clinical assessment of the patient’s medication response. While multiple assessment scores are used in clinical practice (e.g., Rey Auditory Verbal Learning Test (RAVLT) tests, Montreal Cognitive Assessment (MoCA)), we used MMSE assessment scores in our study because it is a widely used tool to assess cognitive function in both routine clinical practice and research settings [22,23]. The max score for MMSE is 30 points, with ranges from 20 to 24 indicating mild dementia; 13 to 20 indicating moderate dementia, and less than 12 indicating severe dementia [24]. We calculated the difference between MMSE in the current visit and the previous visit to measure the rate of progression of

Alzheimer’s. A discount rate gamma, 0 ≤ γ ≤ 1 was also introduced to determine the present value of future rewards [25]. We used the discount factor γ =0.3. Our total discounted return is represented by:

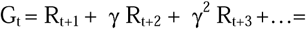

### 2.3. Policies

The policy is a map from state to action. It maps an action to every possible state in the system. In other words, it can be described as a possible strategy an agent uses in each state to get rewards and it is defined by probability. For example, if an agent uses an action a_1_ on state s_1_ and a_2_ on state s_2,_ and so on, it can be considered a policy of the agent. On the state action map, for state s_1_, a_1_ has the highest probability value and for state s_2,_ a_2_ has the highest probability value. There are many possible policies as different actions can be used for the same states; however, one policy will yield the maximum reward.

#### 2.3.1. Optimal policy learned by RL learning

We generated policies using two different RL methods - model-free Q learning and model-based Policy Iteration. Model-based methods rely on planning and transition probabilities, while model-free methods rely on learning or experience [25].

##### • Policy iteration

First, we compute the state-value function v(s) for an arbitrary policy □. Value function, v(s) is a function that estimates future rewards on a given state when performing a particular action based on transition probability. The transition probability is the probability of transitioning from one state, s, to another state, s’ after a certain action is applied. This is called policy evaluation. After computing the value function for a policy, we check if there is a particular action that gives a better value for that state. This is repeated until a better policy is found and is called policy improvement. We repeat these evaluation and improvement cycles until we find out the optimum policy.

##### ⍰ Q Learning

We used this off-policy temporal difference algorithm to create more variety for optimal policies. Q-learning uses Q-value from a Q-table to find the best actions for each state. The Q-value is an estimation of how good an action is at a particular state. The Q table is an m*n matrix where m is the number of states and n is the number of actions. An agent applies an action at a particular state and updates the q-table with the reward it receives for that state-action combination. Then the agent applies different actions for the same state. Through numerous repetitions, the best action for each state is picked and the Q-table becomes stable. The speed at which Q-table is updated is dependent on a parameter alpha, 0 ≤ α <, 1 the learning rate. We set our alpha to 0.05 so that the Q-table converges after enough trials. It is different from policy iteration because it gives an optimal policy independent of the policy being followed. In other words, it is not dependent on transition probability derived from the data set.

#### 2.3.2. Clinicians’ policy by a data-driven approach

We used transition probability to find the clinician’s policy from the data. We followed an approach similar to Policy Iteration. We used policy evaluation and policy improvement process just once based on the existing transition probability from the data and made the resultant policy as the clinician’s policy. Since the policy is totally based on the data, we can safely assume it is very close to the real clinician’s policy.

#### 2.3.3. Other policies

We also created zero policy and random policy to compare them with our RL-based and clinician policies. Zero policy implies that in each state no drugs are applied as actions and random policy implies that random drugs are applied as actions without assessing the patient’s condition.

### 2.4. Experiment Design and Evaluation

#### 2.4.1. Evaluation and comparison

We used offline evaluation to estimate the value of target policies (policies being learned) based on a behavior policy (policy used to generate behavior) [25] derived from the offline log data. It is very useful in settings where online interaction involves high risks and costs (e.g., medication recommendation systems) [26]. We used importance sampling (IS), commonly used off-policy evaluations, to estimate expected values under one distribution given samples from another [25]. It estimates the value of a target policy from behavior policy derived from the data by re-weighing states based on the frequency of their occurrence [27]. In our study, we used stepwise weighted importance sampling (step-WIS) which is the most practical point estimator among the importance of sampling techniques because of its low variance [15] [28] and error [29].

#### 2.4.2. Tests

⍰ <u>Test 1</u>: The first test evaluated the impact of data size in generating policies from AD data in order to create a policy with a higher rate of accuracy and closest to the clinician’s policy. We split 60%/20%/20% for training, validation, and testing. With the training set, we further divided it into four scenarios relating to different data sizes (e.g., 100%, 80%, 50%, 30%) to feed the models. All training groups were trained 50 times to generate an optimal policy. We repeated this cycle 100 times to eliminate any potential bias in our final reward. A total of 13 states and 6 actions were used for this test.

⍰ <u>Test 2</u>: The second test evaluated how the proposed work will perform over the different patient cohorts (e.g., patients with different concomitant diseases). We separated the data into five groups based on the disease diagnosis: Alzheimer’s only (9 states, 6 actions), Hypertension and Alzheimer’s (10 states, 6 actions), Depression and Alzheimer’s (9 states, 6 actions), and Hypertension, Alzheimer’s, and Depression (10 states, 6 actions). Hypertension and Depression were the two most prevalent concomitant diseases in the population in the data. Also, depression is one of the most prevalent psychiatric conditions in AD patients [30–32]. We then followed the same splitting method used in Test 1. We also wanted to check how different RL’s medicine prediction is for different states compared to the clinician’s prediction.

⍰ <u>Test 3</u>: For our final test, we wanted to learn how our proposed Q-learning model performed over different learning rates, α. This test was to confirm our Q-learning was robust enough to learn the real clinician’s policy. We used our already existing Alzheimer’s disease-only cohort and compared the results for alphas from 0.1 to 0.9. We then followed the same splitting method used in Test 1.

⍰ <u>Test 4</u>: For our final test, we wanted to learn how our proposed Q-learning model performed over the different number of states while keeping the data constant. We changed the total number of discrete states given by a decision tree based on the number of samples. For example, for whole data, we got 13 states when we used leaf nodes of a decision tree that had more than 50 samples and 9 states with leaf nodes that had more than 200 samples and compared the results (Fig. 7). We then followed the same splitting method used in Test 1.

**Fig. 3.**
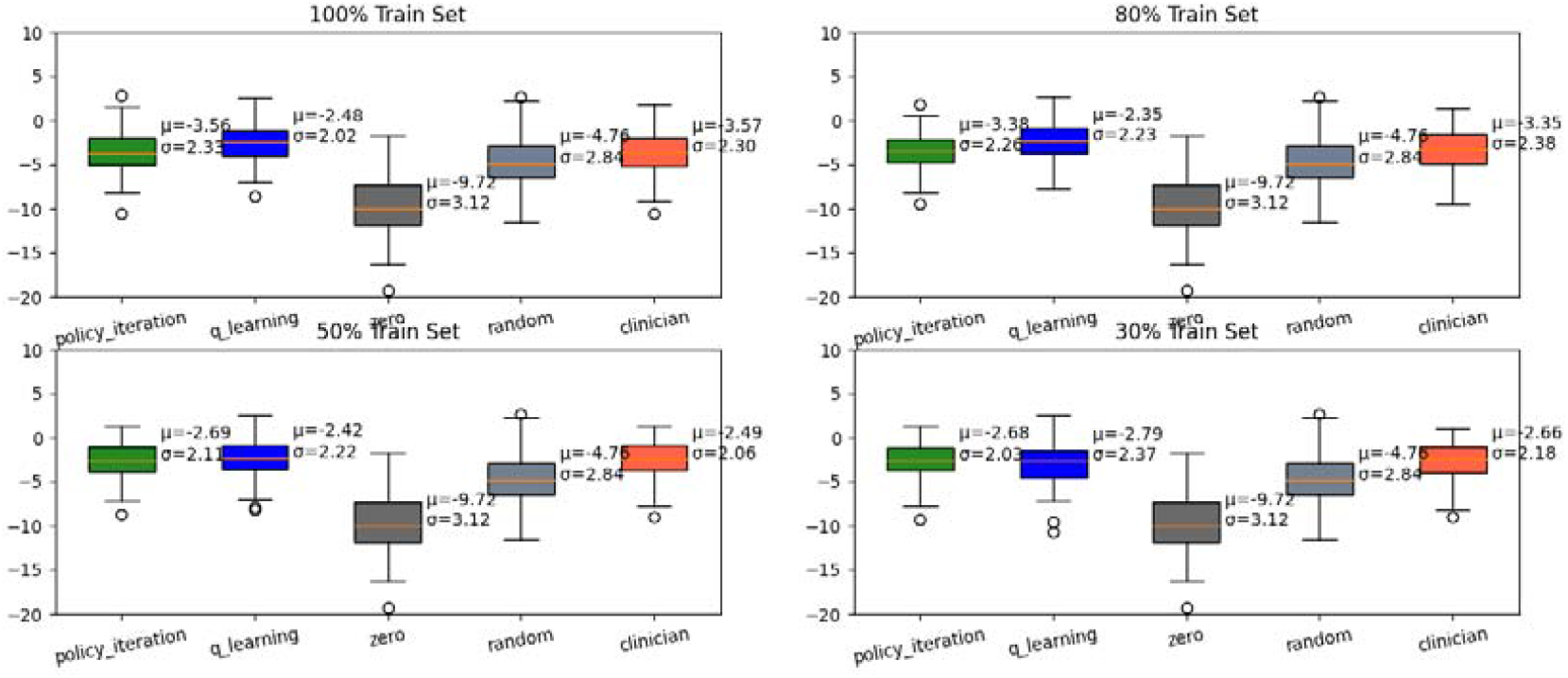
Comparison of rewards represented by MMSE score (y-axis) for different-sized data for all policies. Policy Iteration and Q-learning are the optimal policies, and the Clinician policy is derived from the data.

**Fig. 4.**
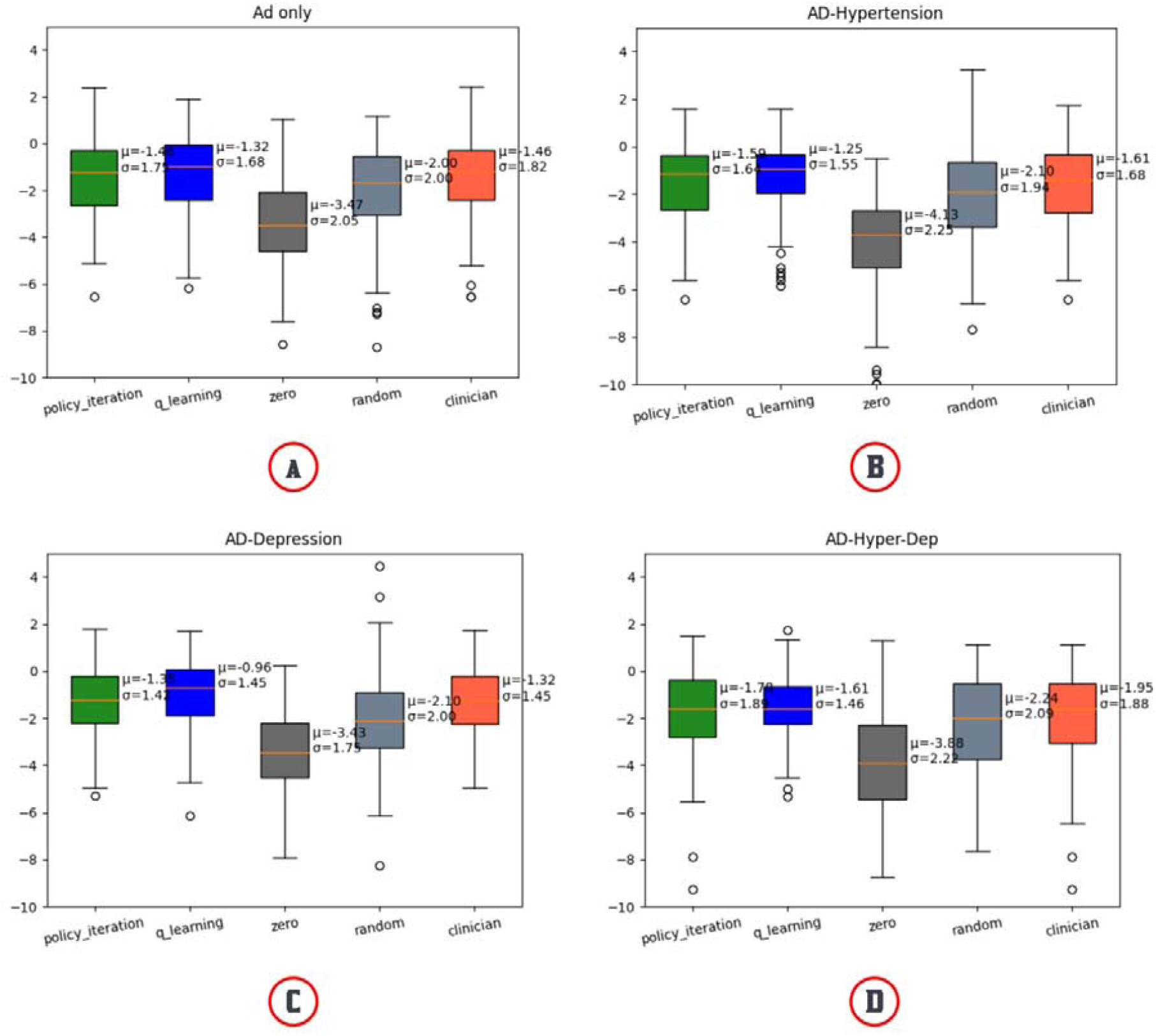
Comparison of different optimal policies (Policy Iteration & Q-learning) and the Clinician’s Policy for different concomitant disease cohorts. (A) Comparison for AD-only patients, (B) Comparison for AD patients with concomitant disease hypertension only, (C) Comparison for AD patients with depression, and (D) Comparison for AD patients with hypertension and depression

**Fig. 5.**
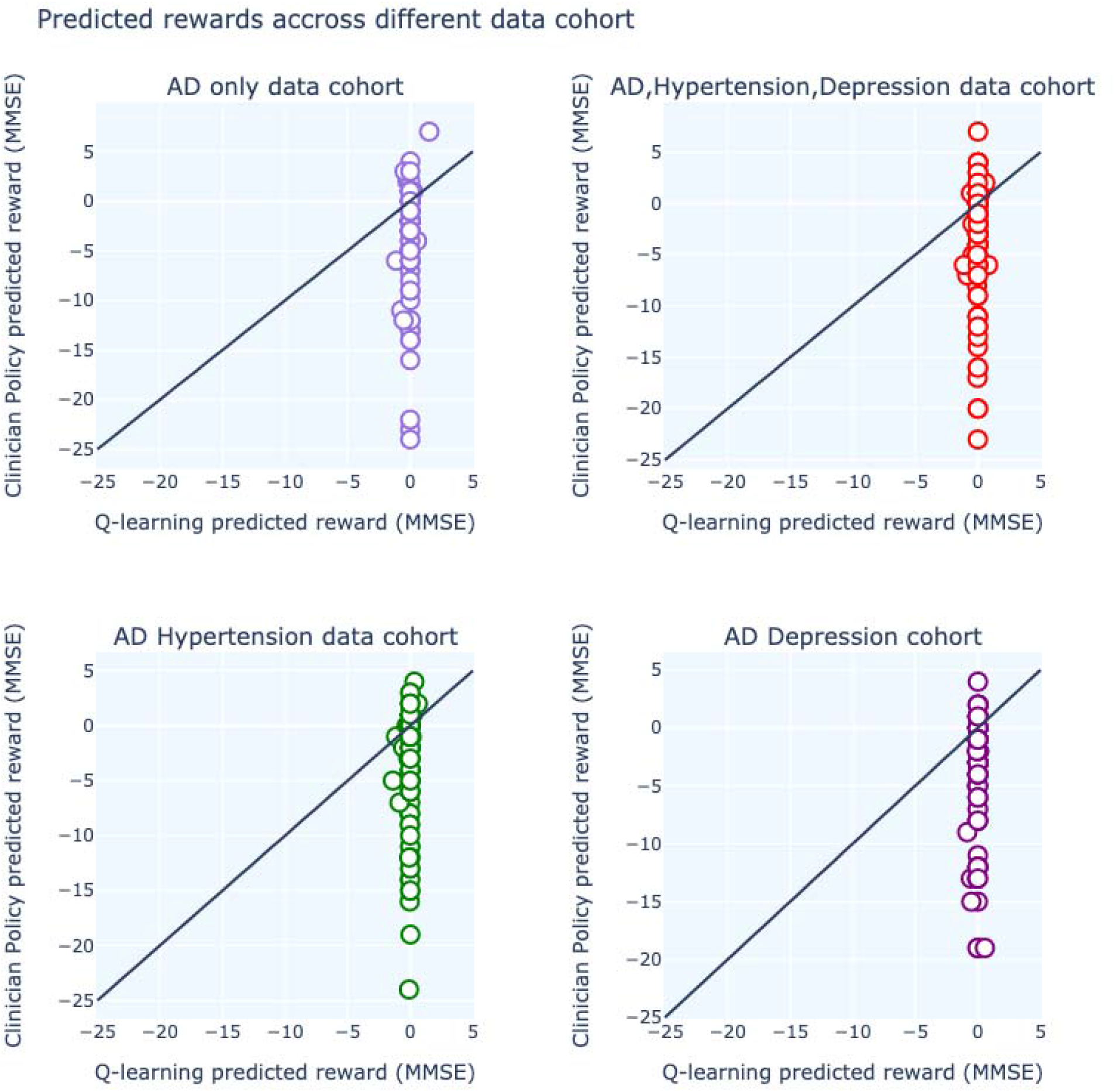
Comparison of reward prediction for different states between Q-learning and clinician’s policy for different data cohorts.

**Fig. 6.**
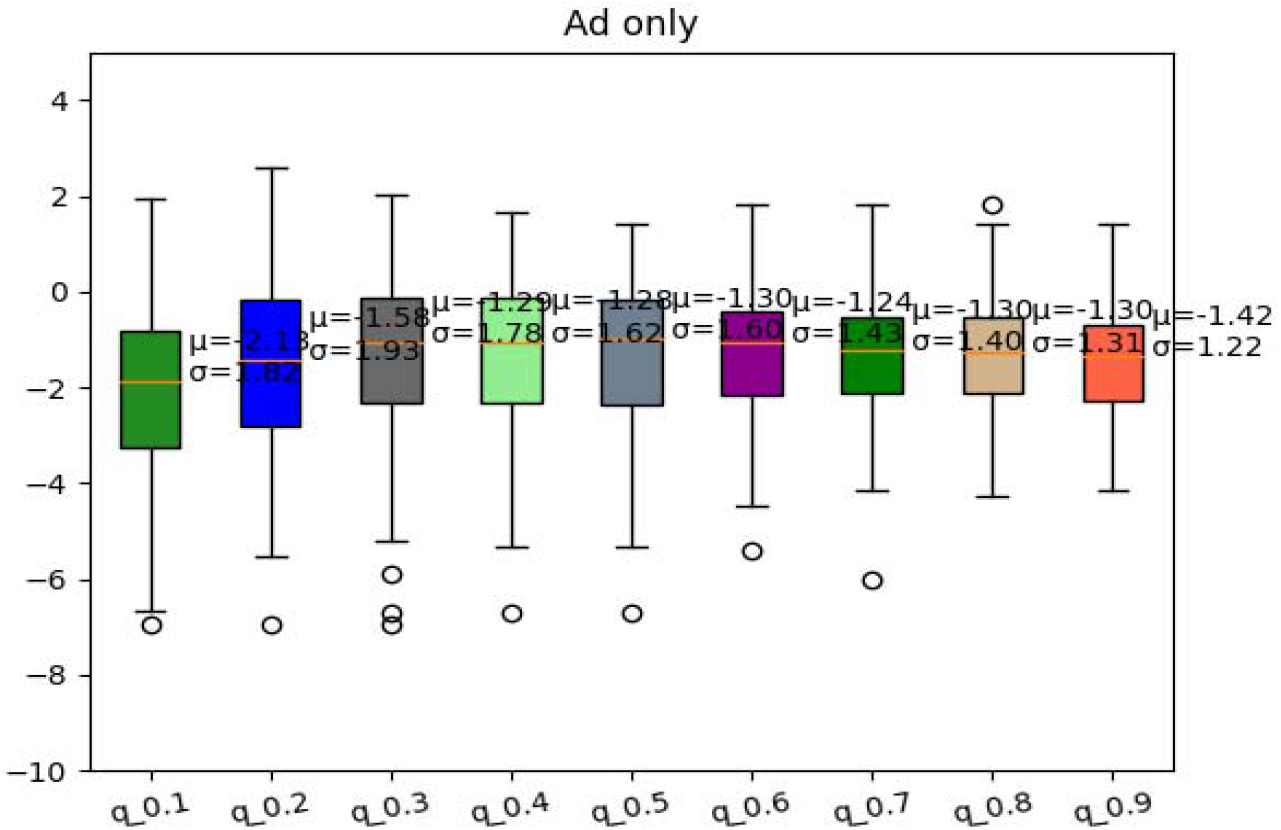
Comparison of Q-learning policy for different learning rates for AD-only cohort. The learning rate is from 0.1 to 0.9.

**Fig. 7.**
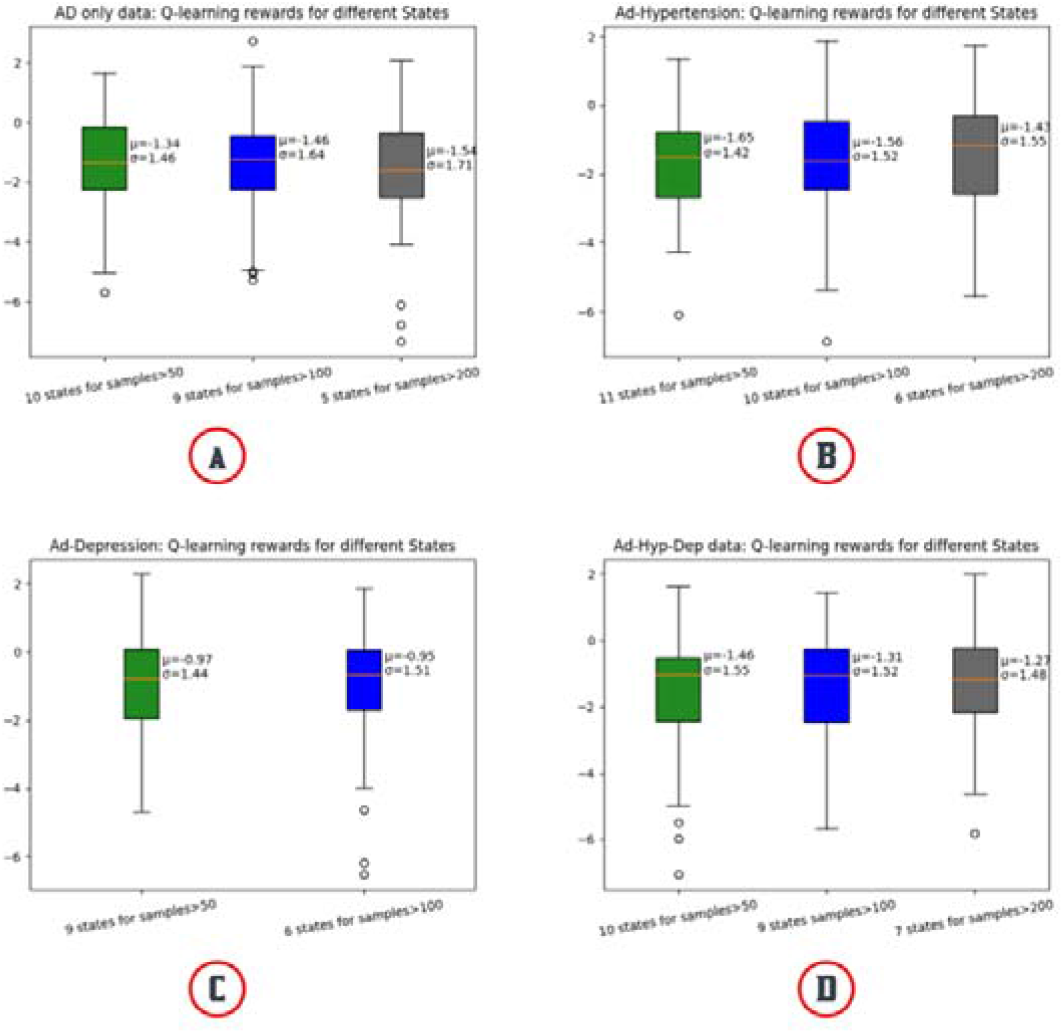
Comparison of Q-learning policy for the different number of states for different data cohorts. The number of states is based on the number of samples on the leaf node of a decision tree.

## 3. Results

### 3.1. Test 1

Test one revealed that appropriate data size resulted in RL performance comparable to the clinician’s performance. Smaller data samples displayed worse results in policy iteration and q-learning than the clinician’s policy, but as data size increased, policy iteration and q-learning performed better than the clinician’s policy. For example, the 30% train set had a lower policy iteration [mean=-2.68] and q-learning [mean=-2.79] than the clinician’s policy [mean=-2.66] compared to the 100% train set where both policy iteration [mean=-3.56] and q-learning [mean=-2.48] showed better results compared to clinician’s policy [mean= -3.57]. Overall, optimal policy consistently performed better than zero policy (where no drugs were applied) and random policy (random drugs were applied). Zero policy repeatedly yielded the lowest mean reward of -9.72 and the lowest single reward (∼ -15) and the random policy was slightly better than the zero policy [mean = -4.76].

The suggestions made by both optimal policies and clinicians’ policies are somewhat similar (Table 3). Both policy iteration and Q-learning start off by recommending no drugs when patients are in the first state whereas the clinicians recommend memantine. In state 11, all the policies recommend hypertension whereas, in state 12, the recommendation by each policy is totally different. In state 6, both optimal policies recommend Hypertension whereas clinicians recommend memantine. Different actions recommendation for each state for AD-Hypertension-Depression Cohort can be found in Supplement Figure 2.

**Table 3.**
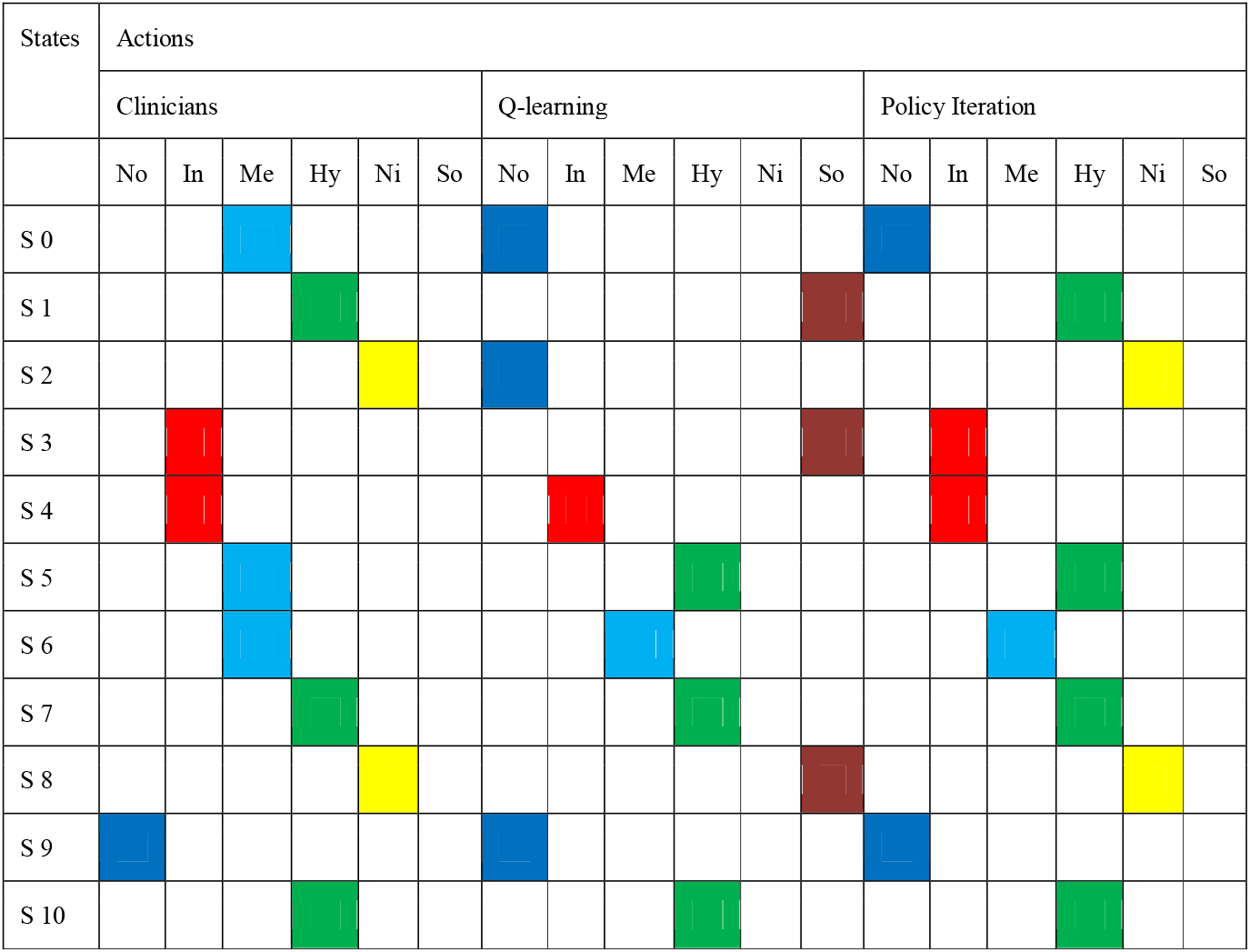
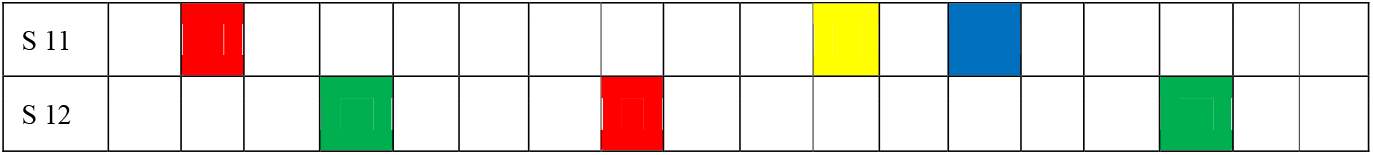
Comparison of Action recommendations for Policy Iteration, Q-learning, and Clinician’s policy for whole Data. In action columns, No is no drugs, In is inhibitors, Me is memantine, Hy is hypertension drugs, Ni is the combination of Memantine and inhibitors and So is supplements/other drugs. The actions are represented by different colors.

### 3.2. Test 2

We noticed that the model is comparable with the clinician’s policy when data is split around AD itself. Since hypertension and depression are frequently seen in AD patients and our actions are mainly the medication for AD, policy iteration showed better results in all three cohorts than the clinician’s policy (Fig. 4). We also compared the MMSE score prediction by Q-learning’s decision-making and Clinician’s policy-making throughout every data cohort and concluded that Q-learning’s rewards are more coherent than clinician’s. For all the data cohorts, Q-learning’s reward predictions are scattered around 0 (lower negative values) whereas clinician reward predictions are scattered around higher negative values rewards (Fig. 5). Q-learning’s reward prediction for whole data can be found in Supplement Figure 3. Predicted reward for each patient for both Q-learning and Clinician’s policy can be found in Supplement Figure 1.

### 3.3. Test 3

We also confirmed the Q-learning policy is not always better with high learning rate (alpha) values. There’s a general trend of increasing rewards from a learning rate of 0.1 to 0.4. Then, the reward is stable from the alpha value of 0.3 to around 0.8 with a mean from -1.28 to -1.30 and then it decreases at 0.9 with a reward of -1.42 (Fig. 6).

### 3.4. Test 4

We did not find any concrete connection between changing the number of states and mean reward prediction (Fig. 7). In the AD-only data cohort, 10 states from leaf nodes with a sample size greater than 50 predicted better mean reward [mean=-1.34] compared to 7 states from leaf nodes with a sample size greater than 200 [mean=-1.54] and 9 states from leaf nodes with a sample size greater than 100 [mean=-1.46]. In the AD-depression data cohort, 9 states from leaf nodes with a sample size greater than 50 predicted worse mean reward [mean=-0.97] compared to 6 states from leaf nodes with a sample size greater than 100 [mean=-0.95] (Fig. 7). This analysis for whole data cohort can be found in Supplement Figure 4.

## 4. Discussion

Our current study proposed an RL-based model to investigate the optimal AD treatment regimen plan based on the EHR. We adopted two RL methods - model-free Q learning and model-based Policy Iteration - to generate the regimen plans. In comparison to the policy (i.e., treatment regimen plan) learned simply from the existing data (i.e., clinician’s policy based on transition probability-based method), the experiments displayed RL models that can optimize the treatment regimen for AD given sufficient patient data as suggested by previous studies with Parkinson’s [15] and sepsis [16]. However, our current study has notable differences compared to those studies. First, we argued that the AI models can only estimate an ideal physician, which is not comparable to a real clinician’s policy, unlike previous studies that strongly suggest AI-based policies can outperform physician policy [15]. Secondly, in previous studies, all the policies were generated based on the on-policy methods (e.g., SARSA and value interaction [15], [16]), which consider the target policy to be identical to the behavior policy. This is problematic in an offline setting because our target policy is very different from the behavior policy as we are using different actions for different states inorder to find an optimal action for a particular state. As a response, we conducted an evaluation that fairly compared the offline model-free models (i.e., Q learning) with the behavior policy. Lastly, we incorporated the importance of data volume to learn an ideal model for real-world implementation in addition to focusing on the RL model performance. Experiments on different data cohorts revealed better RL-based model performance in larger data cohorts. Our experiment showed a harmonization should be achieved between the data and method to generate an optimal policy. In our study, we found the optimal policy by repeating experiments with the training and validation data 50 times. For generalizability, we used 100 bootstrap samples of training and testing data on the resulting optimal policy to find our final reward. This study provided a robust guide for treatment plan learning and has adaptable potential in guiding the treatment of AD patients for junior physicians.

Our results were promising and demonstrated high potential for RL-based models to learn real clinician’s policies; however, there are a few limitations to address. First, we could not obtain definitive results from the latest offline reinforcement learning algorithm, like Conservative Q-Learning (CQL), as it consistently predicted supplements as the optimal action. This is due to the high number of cases where supplements (N=4573) (e.g., vitamin and sleeping medication) were prescribed. This contrast with previous studies examining Parkinson’s Disease (PD) which did not have higher rates of prescribed supplements (N=442) compared to PD medications (Levodopa=1157 and Dopamine agonist=447). There is a lot of potentials to perform this study by using the latest RL algorithms like CQL if evenly distributed medication data is collected in the future.

A second limitation lies in the accuracy of calculating disease progression with only cognitive assessment data. We could not incorporate neuroimaging and other biomarkers data as these were not available. Although there’s no exact way to measure the progression of AD, neuroimaging has been widely used to diagnose AD and monitor disease progression [33]. Due to the unavailability of such data, we had to rely on commonly used cognitive tests like MMSE, ADAS, and CDRSB. A more in-depth study can be performed by incorporating other measures (e.g., mobility) or biomarkers (e.g., amyloid-beta and tau).

Thirdly, we also encountered a lot of negative values in our reward. It could be the result of the small data set, inconsistent data entry for MMSE scores for patients, and the high number of missing values in the record. We tried to minimize the missing values by filling the missing spot with the data from previous visits. The rewards would be much better if accurate MMSE scores were present for each visit for all the patients.

Lastly, there was not an active RL environment to test our algorithms as it is almost impossible to have an active testing environment for medical patients. Off-policy RL algorithms are only successful when they receive direct feedback from an active environment (e.g. a video game). With a proper dataset with evenly distributed medications and fewer missing values, we could use highly effective offline RL algorithms like CQL in the future to avoid this problem [34].

## Supporting information

revised_paper_v24.docx

## Data Availability

All data produced are available online at https://adni.loni.usc.edu/data-samples/access-data/

## Acknowledgment

This work is supported by a grant from the National Institute of Health (NIH) NIGMS (R00GM135488). Data collection and sharing for this project was funded by the Alzheimer’s Disease Neuroimaging Initiative (ADNI) (National Institutes of Health Grant U01 AG024904) and DOD ADNI (Department of Defense award number W81XWH-12-2-0012). ADNI is funded by the National Institute on Aging, the National Institute of Biomedical Imaging and Bioengineering, and through generous contributions from the following: AbbVie, Alzheimer’s Association; Alzheimer’s Drug Discovery Foundation; Araclon Biotech; BioClinica, Inc.; Biogen; Bristol-Myers Squibb Company; CereSpir, Inc.; Cogstate; Eisai Inc.; Elan Pharmaceuticals, Inc.; Eli Lilly and Company; EuroImmun; F. Hoffmann-La Roche Ltd and its affiliated company Genentech, Inc.; Fujirebio; GE Healthcare; IXICO Ltd.; Janssen Alzheimer Immunotherapy Research & Development, LLC.; Johnson & Johnson Pharmaceutical Research & Development LLC.; Lumosity; Lundbeck; Merck & Co., Inc.; Meso Scale Diagnostics, LLC.; NeuroRx Research; Neurotrack Technologies; Novartis Pharmaceuticals Corporation; Pfizer Inc.; Piramal Imaging; Servier; Takeda Pharmaceutical Company; and Transition Therapeutics. The Canadian Institutes of Health Research is providing funds to support ADNI clinical sites in Canada. Private sector contributions are facilitated by the Foundation for the National Institutes of Health (www.fnih.org). The grantee organization is the Northern California Institute for Research and Education, and the study is coordinated by the Alzheimer’s Therapeutic Research Institute at the University of Southern California. ADNI data are disseminated by the Laboratory for Neuro Imaging at the University of Southern California. We appreciate the valuable insight from Dr. David S Knopman to help us improve the manuscript.

## Data and Code Availability

Data used in the preparation of this article were obtained from the Alzheimer’s Disease Neuroimaging Initiative (ADNI) database (adni.loni.usc.edu). As such, the investigators within the ADNI contributed to the design and implementation of ADNI and/or provided data but did not participate in the analysis or writing of this report. A complete listing of ADNI investigators can be found at: http://adni.loni.usc.edu/wp-content/uploads/how_to_apply/ADNI_Acknowledgement_List.pdf. The data can be accessed via https://adni.loni.usc.edu/data-samples/access-data/. The used code is publicly available at https://github.com/bioIKEA/Treatment_optimization_AD

## Notes

### Competing Interest Statement

The authors have declared no competing interest.

### Funding Statement

This study was funded by a grant from the National Institute of Health (NIH) NIGMS (R00GM135488).

