## Supplementary material for "Using Artificial Intelligence to Learn Optimal Regimen Plan for Alzheimer’s Disease": revised_paper_v24.docx

***Supplement 1***

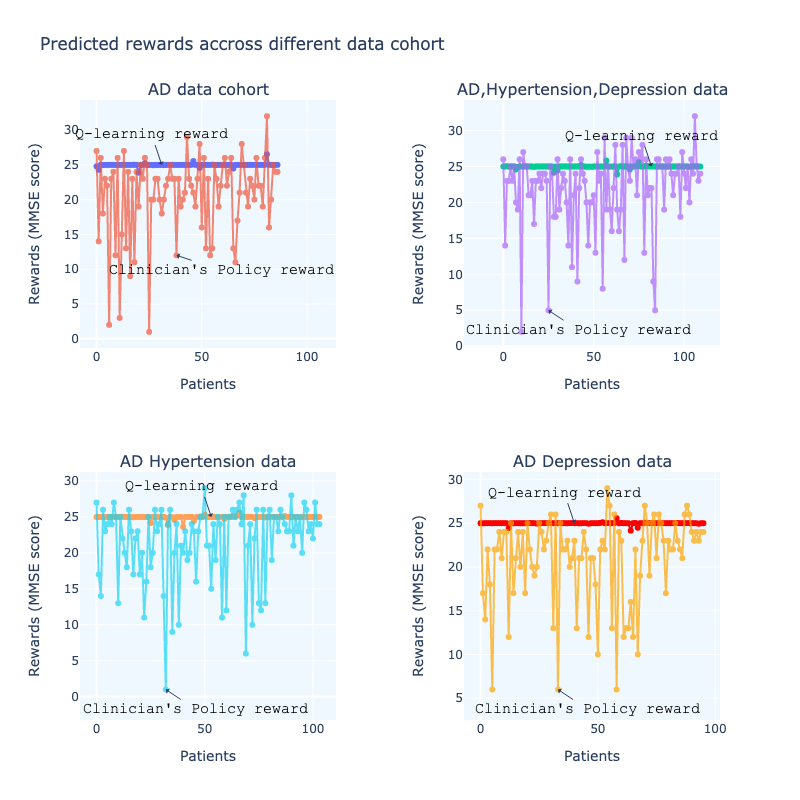

**Supplement Figure 1.** Rewards predicted by Q-learning and Clinician’s Policy for different Patients

***Supplement 2***

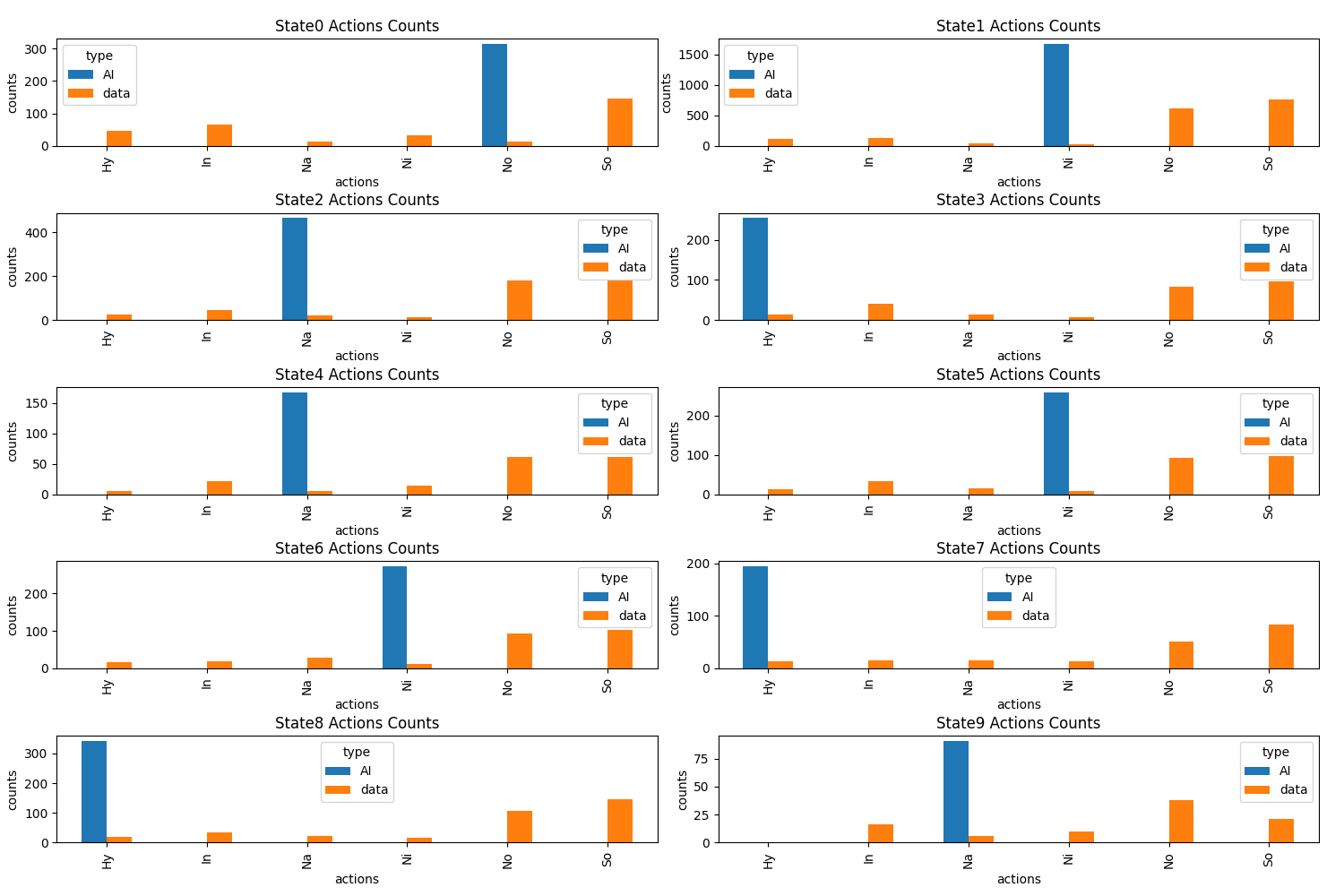

**Supplement Figure 2.** Drugs (actions) recommendation counts for each state by Q-learning and Clinician’s policy for AD-Hypertension-Depression Cohort. In X-axis, No is no drugs, In is inhibitors, Me is memantine, Hy is hypertension drugs, Ni is the combination of Memantine and inhibitors and So is supplements/other drugs.

***Supplement 3***

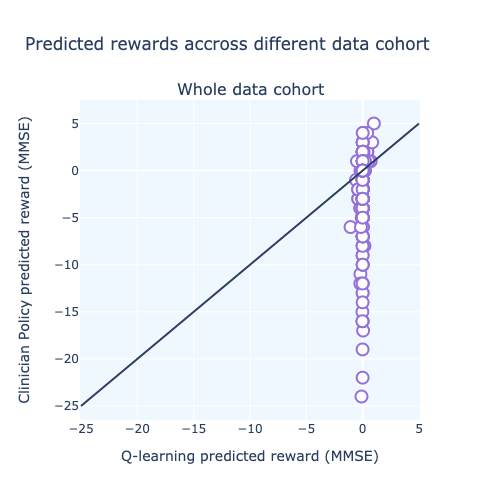

**Supplement Figure 3.** Comparison of reward prediction for different states between Q-learning and clinician’s policy for whole data

***Supplement 4***

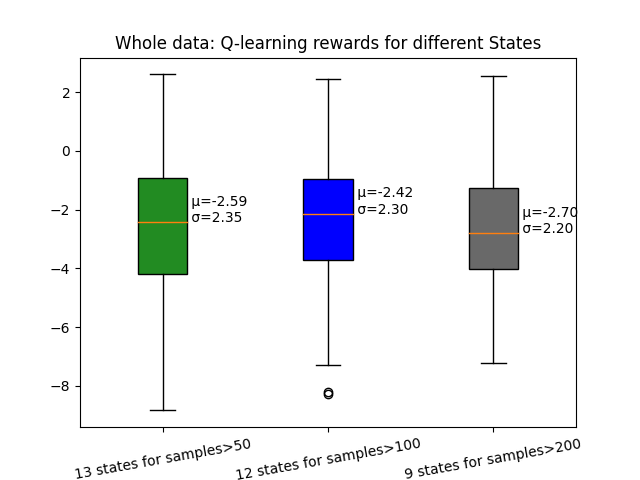

**Supplement Figure 4.** Comparison of Q-learning policy for the different number of states for whole data. The number of states is based on the number of samples on the leaf node of a decision tree.
